# Clinical effectiveness of convalescent plasma in hospitalized patients with COVID-19: a systematic review and meta-analysis

**DOI:** 10.1101/2021.01.16.21249956

**Authors:** Roberto Ariel Abeldaño Zuñiga, Ruth Ana María González Villoria, María Vanesa Elizondo, Anel Yaneli Nicolás Osorio, Silvia Mercedes Coca

## Abstract

Given the variability of previously reported results, this systematic review aims to determine the clinical effectiveness of convalescent plasma employed in the treatment of hospitalized patients with diagnosis of COVID-19.

We conducted a systematic review of controlled clinical trials assessing treatment with convalescent plasma for hospitalized patients with a diagnosis of SARS-CoV-2 infection. The outcomes were mortality, clinical improvement, and ventilation requirement.

A total of 50 studies were retrieved from the databases. Four articles were finally included in the data extraction, qualitative and quantitative synthesis of results. The meta-analysis suggests that there is no benefit of convalescent plasma compared to standard care or placebo in the reduction of the overall mortality and in the ventilation requirement; but there could be a benefit for the clinical improvement in patients treated with plasma.

We can conclude that the convalescent plasma transfusion cannot reduce the mortality or ventilation requirement in hospitalized patients diagnosed with SARS-CoV-2 infection. More controlled clinical trials conducted with methodologies that ensure a low risk of bias are still needed.

## Introduction

The SARS-CoV-2 virus first detected in Wuhan China has caused a global pandemic (1). What is known about the microorganism is established by genomic analysis as the disease spreads (2,3). The pandemic still represents a global health threat, in mid-January 2021, the total number of COVID-19 cases reported by the World Health Organization (WHO) is close to 100 million, while deaths have exceeded 2 million (4). One year after the onset of the disease, it is known that there are cases with different degrees of severity ranging from asymptomatic cases to critical patients, in whom respiratory failure, septic shock or multi-organ failure occurs, requiring various hospital care and supportive treatment (5). WHO points out that there are more than 200 vaccines under investigation, only some in phase three and four are being distributed currently (6–9).

The distribution of these vaccines is subject to the production, acquisition, storage, and distribution capacities of each country. As of January 2021, 90% of the vaccines produced are concentrated in 9 countries (10). These problems limit a large percentage of the world population to be vaccinated as soon as possible and therefore reach herd protection, this limitation being even greater in low-income countries, which will have to wait a considerable time longer. This is due to the fact that the pre-order manufacturing contracts for vaccines to 13% of the population mainly in the European Union (6). All this implies that people will continue to be infected and will continue to die from this infection in low-income countries. Therefore, to find a treatment that reduces the severity of the disease and reduces the incidence of fatality is still a major public health concern.

On the other hand, in addition to the limitations of access to the vaccine, their efficacy has been established in a very wide range, from 50% to 95% (8,9). Therefore, in countries where there will not be prompt protection of the population, it is necessary to use treatments for the recovery of hospitalized patients when the effectiveness of the vaccines is not as expected.

Among the repurposed treatments, the use of passive immunity has been suggested as an alternative since the beginning of the pandemic (11). The use of convalescent plasma as a treatment against COVID-19 was approved in March 2020 by the FDA (12). The treatment uses the administration of antibodies collected from people recently infected and recovered from COVID-19. The results of the use of plasma are variable, reporting efficacy if its use is in the first sixteen days of illness was associated with an improvement in the first days after treatment, as well as lower requirements for ventilatory support, on the other hand there are studies that show that the disease is in a moderate phase plasma treatment does not show evidence of preventing disease progression (13–19). Given the variability of previously reported results, the present work aims to determine the clinical effectiveness of convalescent plasma employed in the treatment of hospitalized patients with diagnosis of COVID-19.

## Methods

A systematic review was conducted adhering to the PRISMA guidelines for conducting systematic reviews (20). The question in this review was: What is the clinical effectiveness of convalescent plasma employed in the treatment of hospitalized patients with diagnosis of COVID-19? To conduct the review, the PICOS structure was followed according to these points:

- Patients: Adults hospitalized with a diagnosis of SARS-CoV-2 infection.
- Intervention: Treatment with convalescent plasma.
- Comparison: Placebo or standard care.
- Outcomes: Overall mortality, clinical improvement at seven days, clinical improvement at 14 days, clinical improvement at 28 days, duration of ventilation (days), duration of hospitalization (days), virological clearance, and severe adverse events.
- Studies (type of): Clinical trials published in peer-reviewed journals.

The search was carried out in PubMed, Scopus, and Web of Science databases, between November 20th, 2020, and January 9th, 2020. The references of the selected articles were also reviewed for an integral reading to include additional studies not indexed in these databases. The clinicaltrials.gov website was also scanned to obtain potential published reports of registered trials. The search strategies included the following keywords: convalescent plasma, COVID-19, SARS-CoV2, and hospitalized. See the supplemental file for more details on the search strategies.

Studies that met the following criteria were included: I) Controlled clinical trials, II) Studies that included hospitalized patients with SARS-CoV-2 infection, III) Published in 2020 and 2021, IV) Published in English, Chinese, Spanish, or Portuguese. The exclusion criteria were I) Not being a clinical controlled trial, II) Not treating hospitalized patients, and III) Nos using convalescent plasma.

All references were managed with Mendeley® software. The selection of the articles began with the removal of duplicate articles and proceeded with the reading of the title and abstract, carried out independently by reviewers 1, 2, and 3. The final decision in cases of disagreement was based on the criteria of a fourth reviewer. In the second phase, the same reviewers read the full text of the studies to define which would be included for the extraction and synthesis of data. The data were stored in Microsoft Office Excel spreadsheets and organized in an instrument constructed by the authors considering: Characteristics of the study (author, year, country), sample, study design, and characteristics of the results. The risk of bias of the studies was evaluated using the ROB2 tool (21). The included studies were independently assessed by reviewers 1 and 5 (See supplemental file).

The qualitative synthesis was developed following the assessed outcomes: Overall mortality, clinical improvement at seven days, clinical improvement at 14 days, clinical improvement at 28 days, ventilation requirement, hospital stay (days), virological clearance, and severe adverse events.

### Statistical analysis

Meta-analyses of inverse variance were conducted for three outcomes: clinical improvement at day 7, ventilation requirement and overall mortality. Meta-analyses were conducted with Revman v5.4 using pooled fixed effects odds ratios. The significance and the magnitude of heterogeneity across studies were calculated using the Q and I2 statistics. Odds ratios with 95% CIs were plotted for the association between convalescent plasma compared with standard care or placebo. Subgroup analyses were performed to examine differences according to the population (All adults and older adults) in the treatment with convalescent plasma for the ventilation requirement and the overall mortality in patients diagnosed with SARS-Cov-2 infection.

The review protocol was registered on the PROSPERO platform (CRD42020184436).

## Results

Following the described PICOS structure, this systematic review retrieved 50 studies from the databases. After the removal of 4 duplicates, 46 articles were read in title and abstract. Forty-two were eliminated, resulting in 4 articles for full-text reading. Four articles were finally included in the data extraction, qualitative and quantitative synthesis of results (Figure 1).

**Figure 1.**
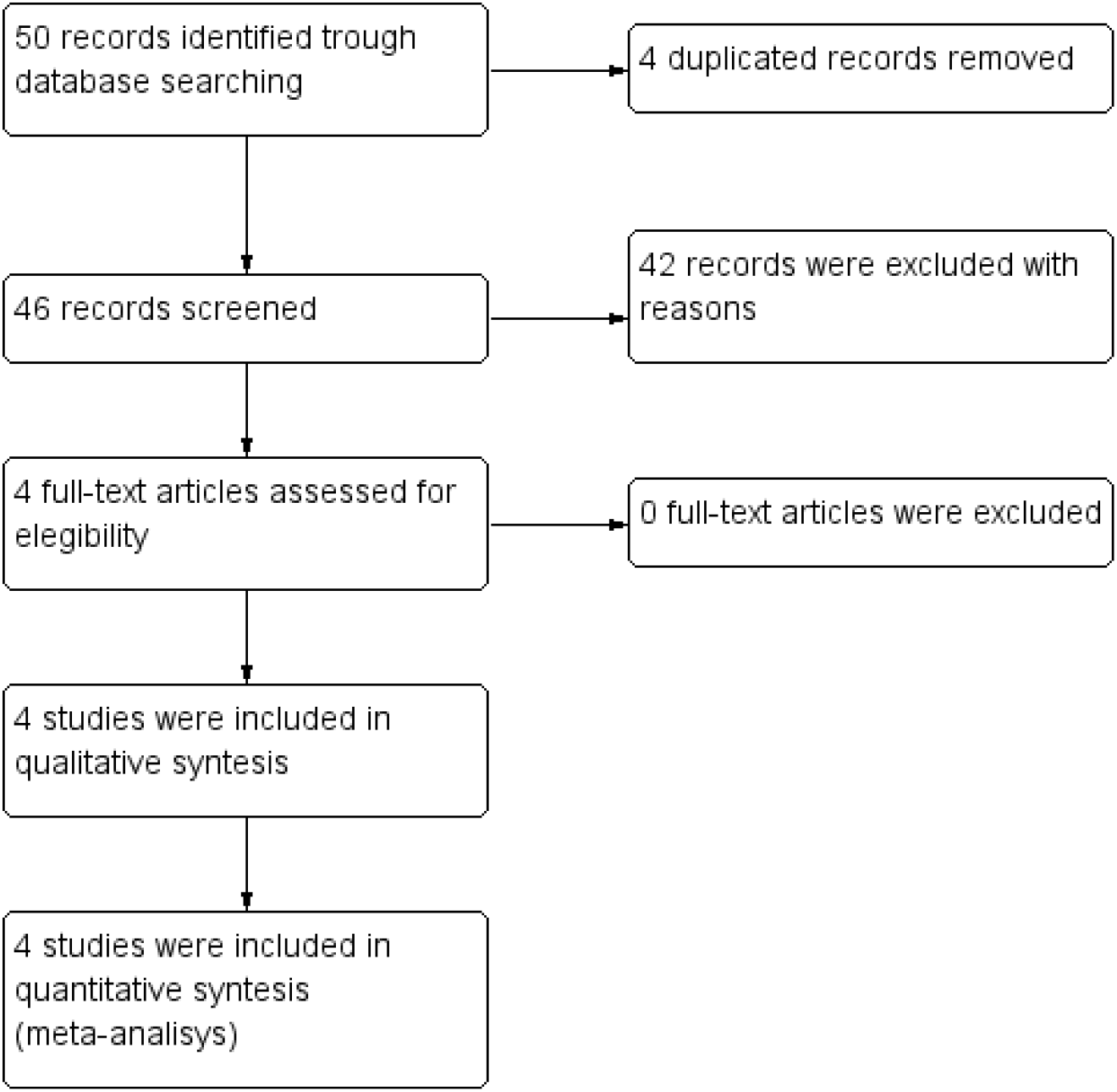
Prisma flowchart of the inclusion process in the systematic review.

The overall risk of bias in the reviewed articles was established at low-risk only in one randomized, open-label clinical trial (17). The remaining three studies were established at high risk of bias due to issues in the randomization process (16,18,19). More details can be seen in the supplemental file. Patient samples ranged from 103 (the study with the fewest patients) to 464 (the study with the most patients); three clinical trial included adult patients (16,18,19), while the fourth study was focused only in older adult patients (17). The retrieved results were: Mortality, clinical improvement at 7, 14, and 28 days (defined by clinical scales), ventilation requirement, the mean duration of hospitalization (in days), virological clearance (by laboratory tests), progression to severe disease and severe adverse events (Table 1).

**Table 1.** Main characteristics of included studies

| Author | Study site | Design | Sample | Intervention | Control | Antiviral treatment | Primary outcomes | Secondary outcomes |
| --- | --- | --- | --- | --- | --- | --- | --- | --- |
| Li (19) | China | Randomized, open label, multicentre, clinical trial | 52 intervention, 51 control | Convalescent plasma 4 to 13 mL/kg of recipient body weight. 96% received a single dose transfusion. | Standard care: symptomatic control | Yes, both groups | Clinical improvement at day 7, 14 and 28 | mortality at day 28, hospital stay, virological clearance, |
| Agarwal (18) | India | Randomized, open label, multicentre, clinical trial | 235 intervention, 229 control | 2 doses of 200 ml convalescent plasma, transfused 24 hours apart | Standard care following institutional protocol | Yes, only the control group. | Progression to severe disease and mortality at day 28 | Clinical improvement at day 7, O2 requirement, respiratory support, ventilation requirement, organ failure, virological clearance, levels of biomarkers, and vasopressor support |
| Abolghasemi (16) | Iran | Non-randomized, multicentre, clinical trial | 115 intervention, 74 control | 1 dose of 500 ml of convalescent plasma. Second dose of 500 ml was administered If the patient did not show any improvement after 24 hours | Standard care | Yes, both groups | Mortality and hospital stay | Ventilation requirement and adverse events |
| Libster (17) | Argentina | Randomized, open label, multicentre, clinical trial | 80 intervention, 80 control | 250 ml of convalescent plasma with IgG titer greater than 1:1000 | 250 ml of placebo (0.9% normal saline) | Not reported | Progression to severe disease | Life threatening respiratory disease, ventilation requirement, admission to ICU, critical systematic illness, multiple organ disfunction, mortality and adverse events |

The four studies reported using convalescent plasma at different dosage. Li (19) reported a 4 to 13 mL/kg of recipient body weight dose, with possibility of receiving a second dose (96% received a single dose transfusion). Agarwal (18) reported 2 doses of 200 ml, transfused 24 hours apart. Abolghasemi (16) reported 1 dose of 500 ml followed by a second dose of 500 ml If the patient did not show any improvement after 24 hours. The study developed by Libster (17) in Argentina was the only one that reported 250 ml of convalescent plasma with IgG titer greater than 1:1000. Also, it is important to highlight that 2 studies used antiviral drugs in both groups (16,19), one study used antiviral treatment in the control group (18), and one study did not reported the use of drugs in any group (17).

### Outcomes assessed

The main outcome assessed by this systematic review was the mortality in hospitalized patients diagnosed with SARS-CoV-2 infection. Two clinical trials assessed the mortality of hospitalized patients at day 28 (18,19), and two studies reported mortality at any time from randomization (16,17) (Table 1).

The clinical improvement was reported by two studies. Li (19) has measured this outcome at days 7,14 and 28 using the National Early Warning Score (NEWS) 2 (22), while Agarwal (18) has measured this outcome at day 7 as the proportion of participants showing resolution of symptoms of fever, shortness of breath, or fatigue. Abolghasemi (16) and Libster (17) did not assessed clinical improvement (Table 1).

The progression to severe disease was assessed by Agarwal and Libster (17,18). The definitions of this outcome differed in both clinical trials: PaO2/FiO2 ratio <100 mm Hg any time within 28 days of enrolment (18); and respiratory rate of 30 bpm or more, SpO2 <93% at ambient air, or both (17) (Table 1).

The virological clearance was reported by two studies (18,19) using different criteria, and all studies reported adverse events identified in patients (16–19). The study published by Li was the only one that assessed subgroups: all patients, patients with severe disease, and patients with life-threatening disease (19). Other assessed outcomes are shown in table 1.

Table 2 shows the main results from the four articles included in the qualitative synthesis. The reduction in overall mortality is supported by Li(19) and Abolghasemi (16), but Agarwal (18) and Libster (17) did not found statistically significant differences.

**Table 2.**
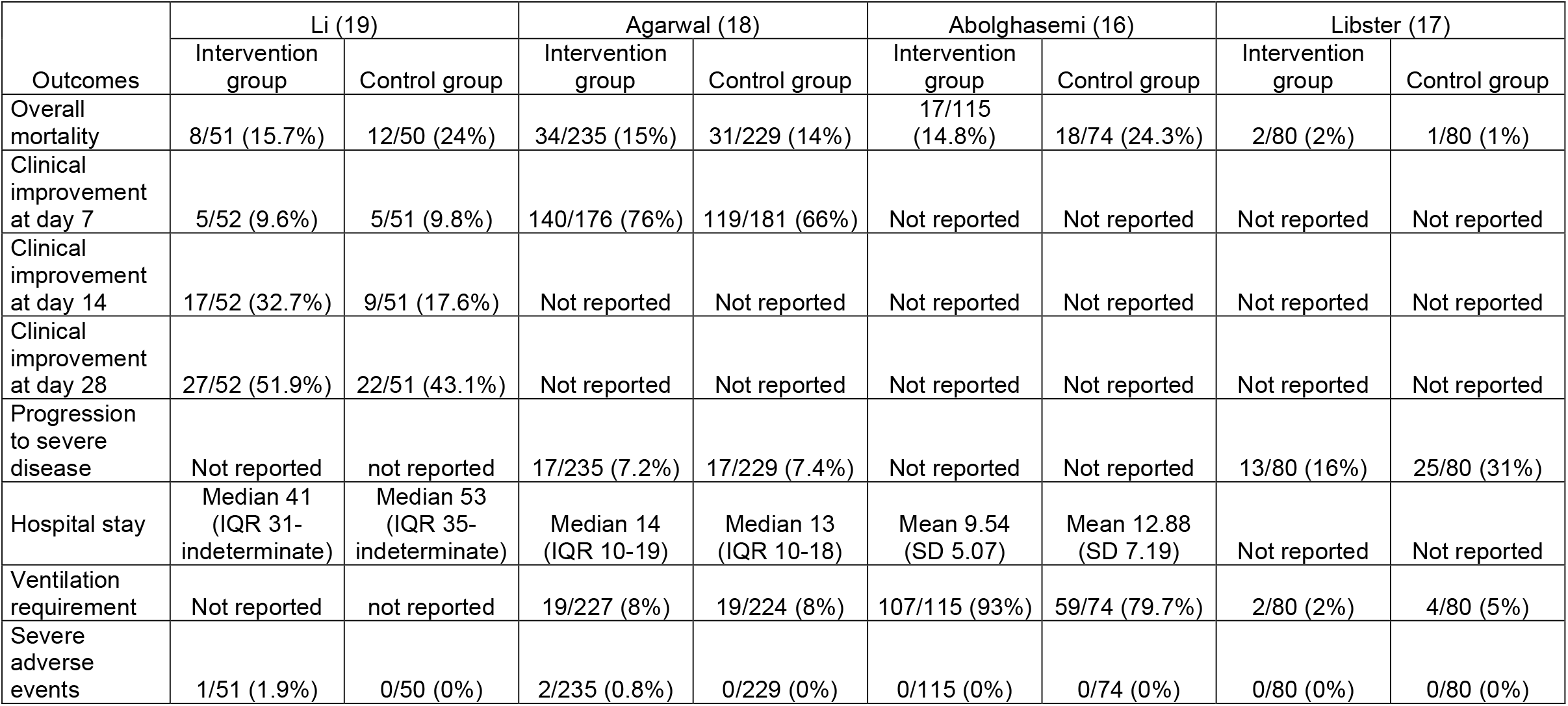
Reported outcomes in the included studies.

| Outcomes | Li (19) |  | Agarwal (18) |  | Abolghasemi (16) |  | Libster (17) |  |
| --- | --- | --- | --- | --- | --- | --- | --- | --- |
|  | Intervention group | Control group | Intervention group | Control group | Intervention group | Control group | Intervention group | Control group |
| Overall mortality | 8/51 (15.7%) | 12/50 (24%) | 34/235 (15%) | 31/229 (14%) | 17/115 (14.8%) | 18/74 (24.3%) | 2/80 (2%) | 1/80 (1%) |
| Clinical improvement at day 7 | 5/52 (9.6%) | 5/51 (9.8%) | 140/176 (76%) | 119/181 (66%) | Not reported | Not reported | Not reported | Not reported |
| Clinical improvement at day 14 | 17/52 (32.7%) | 9/51 (17.6%) | Not reported | Not reported | Not reported | Not reported | Not reported | Not reported |
| Clinical improvement at day 28 | 27/52 (51.9%) | 22/51 (43.1%) | Not reported | Not reported | Not reported | Not reported | Not reported | Not reported |
| Progression to severe disease | Not reported | not reported | 17/235 (7.2%) | 17/229 (7.4%) | Not reported | Not reported | 13/80 (16%) | 25/80 (31%) |
| Hospital stay | Median 41 (IQR 31-indeterminate) | Median 53 (IQR 35-indeterminate) | Median 14 (IQR 10-19) | Median 13 (IQR 10-18) | Mean 9.54 (SD 5.07) | Mean 12.88 (SD 7.19) | Not reported | Not reported |
| Ventilation requirement | Not reported | not reported | 19/227 (8%) | 19/224 (8%) | 107/115 (93%) | 59/74 (79.7%) | 2/80 (2%) | 4/80 (5%) |
| Severe adverse events | 1/51 (1.9%) | 0/50 (0%) | 2/235 (0.8%) | 0/229 (0%) | 0/115 (0%) | 0/74 (0%) | 0/80 (0%) | 0/80 (0%) |

Li and Agarwal conclusions state that the convalescent plasma transfusion did not result in benefit for the intervention groups (18,19), while the studies published by Abolghasemi and Libster stated that convalescent plasma was clinically effective in COVID-19 patients (16,17). Since the conclusions reported by the included studies differed, we decided to conduct the meta-analysis to obtain global estimations for the outcomes of our interest.

### Meta-analysis

The result of two studies was integrated into the fixed-effects meta-analysis for comparing convalescent plasma versus standard care in the clinical improvement of patients diagnosed with SARS-CoV-2 infection (18,19). In this case, convalescent plasma has shown a benefit for patients (OR: 1.86; CI: 1.19-2.91) (Figure 2).

**Figure 2.**
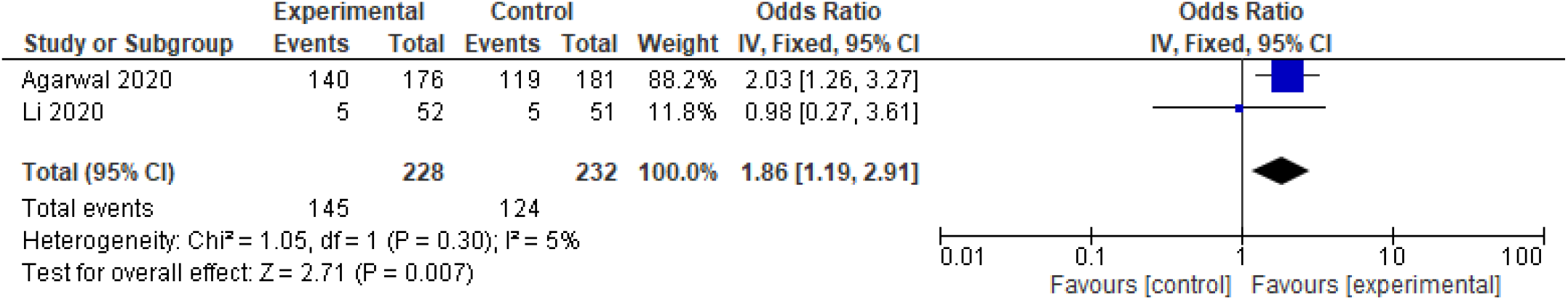
Forest plot of convalescent plasma transfusion for hospitalized patients with SARS-CoV-2 infection Comparison “Convalescent plasma versus standard care”. Outcome: Clinical improvement at day 7.

Three studies reporting ventilation requirement outcomes (16–18) were compared to test the overall effect of convalescent plasma. The results of the random-effects meta-analysis show no association with ventilation requirement (OR:1.34; CI: 0.48-3.72) (Figure 3).

**Figure 3.**
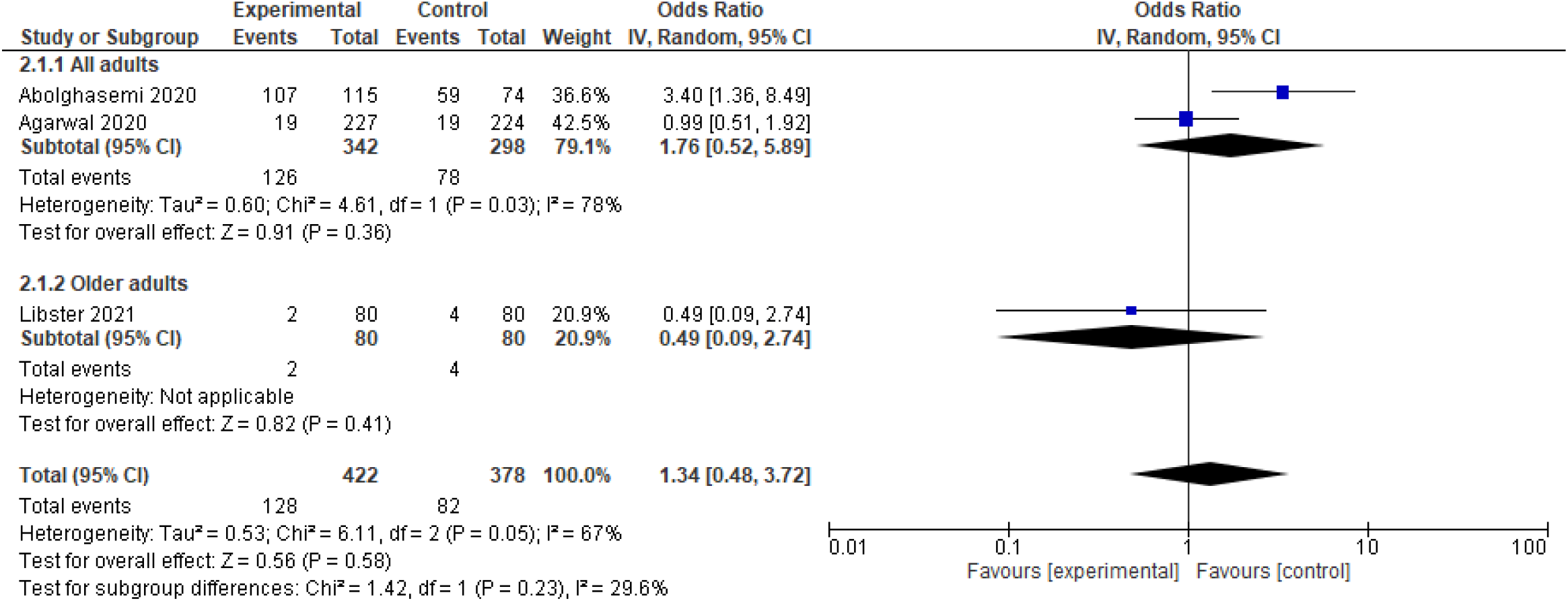
Forest plot of convalescent plasma transfusion for hospitalized patients with SARS-CoV-2 infection Comparison “Convalescent plasma versus standard care”. Outcome: Ventilation requirement.

Finally, the results of 3 studies (16,18,19) were meta-analyzed to establish comparisons on the overall mortality. The meta-analysis of fixed effects suggests no benefits using the convalescent plasma transfusion for reducing the risk of overall mortality (OR: 0.83; CI: 0.56-1.22) (Figure 4).

**Figure 4.**
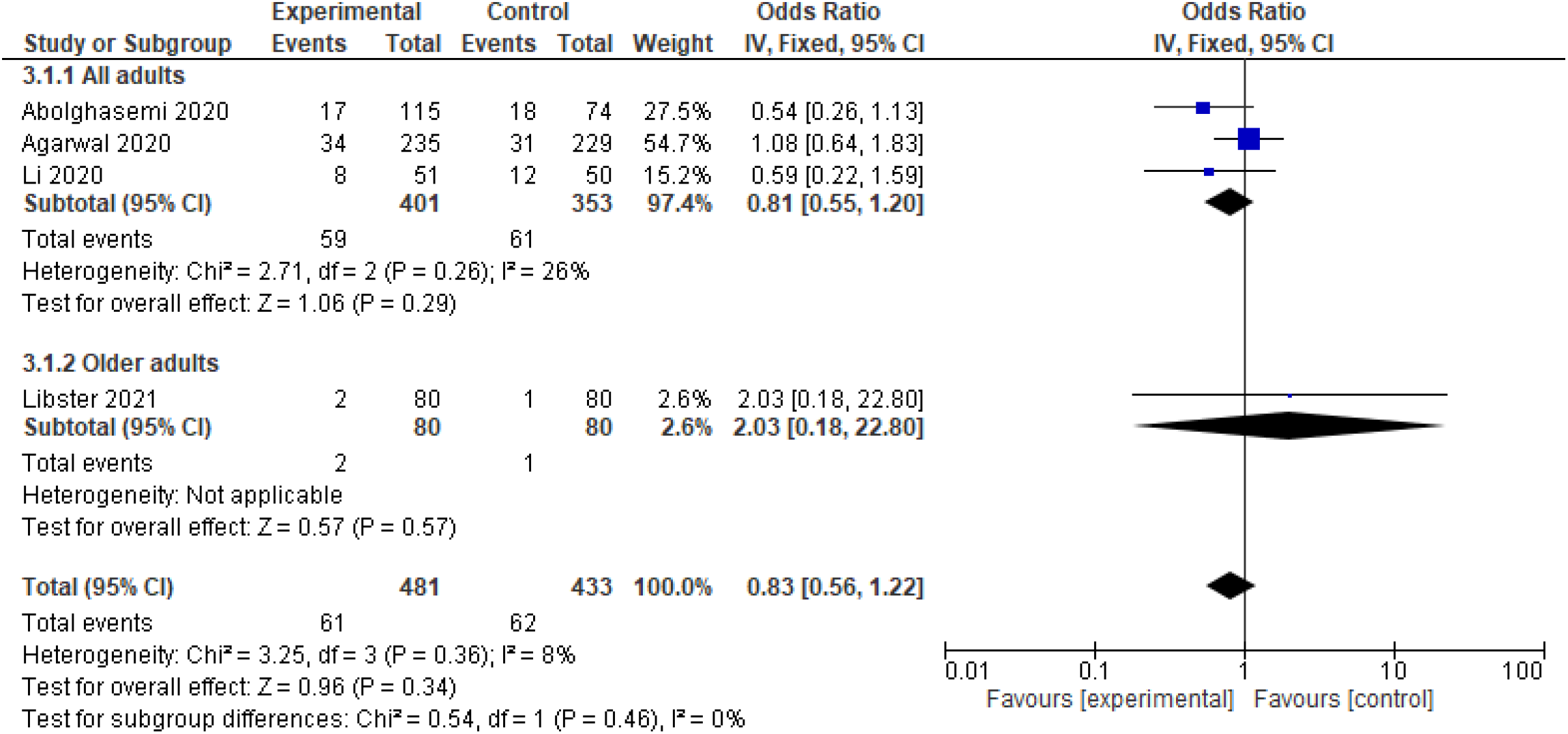
Forest plot of convalescent plasma transfusion for hospitalized patients with SARS-CoV-2 infection Comparison “Convalescent plasma versus standard care”. Outcome: Overall mortality.

## Discussion

This systematic review was focused on of adults hospitalized patients diagnosed with SARS-CoV-2 infection, treated with convalescent plasma transfusion. The studies included in this review were quite heterogeneous regarding the doses of plasma administered, the co-treatment with repositioned antiviral drugs in the experimental group and in the control group, and in the results obtained by each clinical trial. Considering only controlled clinical trials published in peer-reviewed journals, 916 patients were included in hospitals from China, India, Iran, and Argentina.

Regarding the overall quality of the studies, it must be noted that 3 out of 4 clinical trial were considered at high risk of bias due to the lack of blinding. This is a common characteristic of many clinical studies that started to run under emergency conditions due to the persistent health crisis, with recurring waves in some countries in Latin America, and other countries (23).

There are four published systematic reviews on convalescent plasma that have shown that this treatment could reduce the mortality (24–27), but did not included only controlled trials and did not included the last study published by Libster (17). The current systematic review did not showed any benefit on the mortality reduction, consistent with other three published systematic reviews (28–30), while other systematic reviews were focused on other infectious diseases such as Ebola, influenza or SARS (31–33), or other target populations (34).

The use of convalescent plasma was associated with clinical improvement, which is consistent with other previously published study (24,27,29), however, Li (6) has stated that plasma treatment has not been effective in critically ill patients, which suggests that more stratified analysis are needed in primary studies. In addition, the plasma transfusion has not been effective for avoiding the ventilation requirement, as stated previously by Chai (29). All this could suggest that the clinical effects of an earlier transfusion of convalescent plasma should be continue to be assessed in subsequent clinical trials, just like Libster (17) suggests in mildly ill patients in early stages of the infection. In the decade of 1970, one study on hemorrhagic fever in Argentina has shown more effectiveness of convalescent plasma in the first days of the clinical course (35). Still, the optimal dosage and the best time point for the convalescent plasma transfusion need to be determined in well-designed clinical trials (36).

Something that has been sufficiently proven before is that convalescent plasma administration does not have many severe adverse events in transfusion (16– 19,27,31). In contrast, more research is needed on the synergistic effect that plasma could have with other repositioning drugs, as has been demonstrated, for example, with the use of remdesivir as has been published in other studies (19,27,37).

Among the limitations of this study, the rapid generation of new knowledge in times of the pandemic, can potentially affect the timeliness of this review in a few months. The second limitation is the heterogeneity and high risk of bias in the studies. In this review, we chose not to issue recommendations with the GRADE methodology, due to heterogeneity and high risk of bias. Another limitation is that not all studies have used the same dosage of convalescent plasma in infected patients. The fourth limitation that must be considered is regarding the use of antiviral drugs in the control groups or both groups of patients in three out of four clinical trials included in this review.

In times of recurring waves of the COVID-19 pandemic, the analysis of potential treatments proposed for hospitalized patients is still necessary since the procurement and logistics of vaccines are still seen within a complex scenario for many low-income countries. In many low-income countries, people are likely to continue to be infected and to continue to die, where vaccination would occur two to four quarters later partly due to logistical issues, as stated The World Bank (38); so the search for a clinically effective treatment is still a major concern globally.

In the clinicaltrials.gov platform are currently registered dozens of clinical trials that are assessing the treatment with plasma, so the addition of new evidence in the coming months could change the direction of the analyzes in this review.

## Conclusion

The transfusion with convalescent plasma cannot reduce the mortality or ventilation requirement in hospitalized patients diagnosed with SARS-CoV-2 infection. More controlled clinical trials conducted with methodologies that ensure a low risk of bias are still needed.

## Data Availability

All data are available upon request to the corresponding author by mail.

All authors declare that there are no conflicts of interest.

There was no funding for this project.

All data are available upon request to the corresponding author.

## Supplemental file

**Table 1.** Search strategies

|  |  |
| --- | --- |
| 1 | COVID-19 |
| 2 | SARS-CoV-2 |
| 3 | 1 OR 2 |
| 4 | plasma |
| 5 | convalescent<br>plasma |
| 6 | 4 OR 5 |
| 7 | 3 AND 6 |
| FILTER | Clinical Trial |

**Figure 1.**
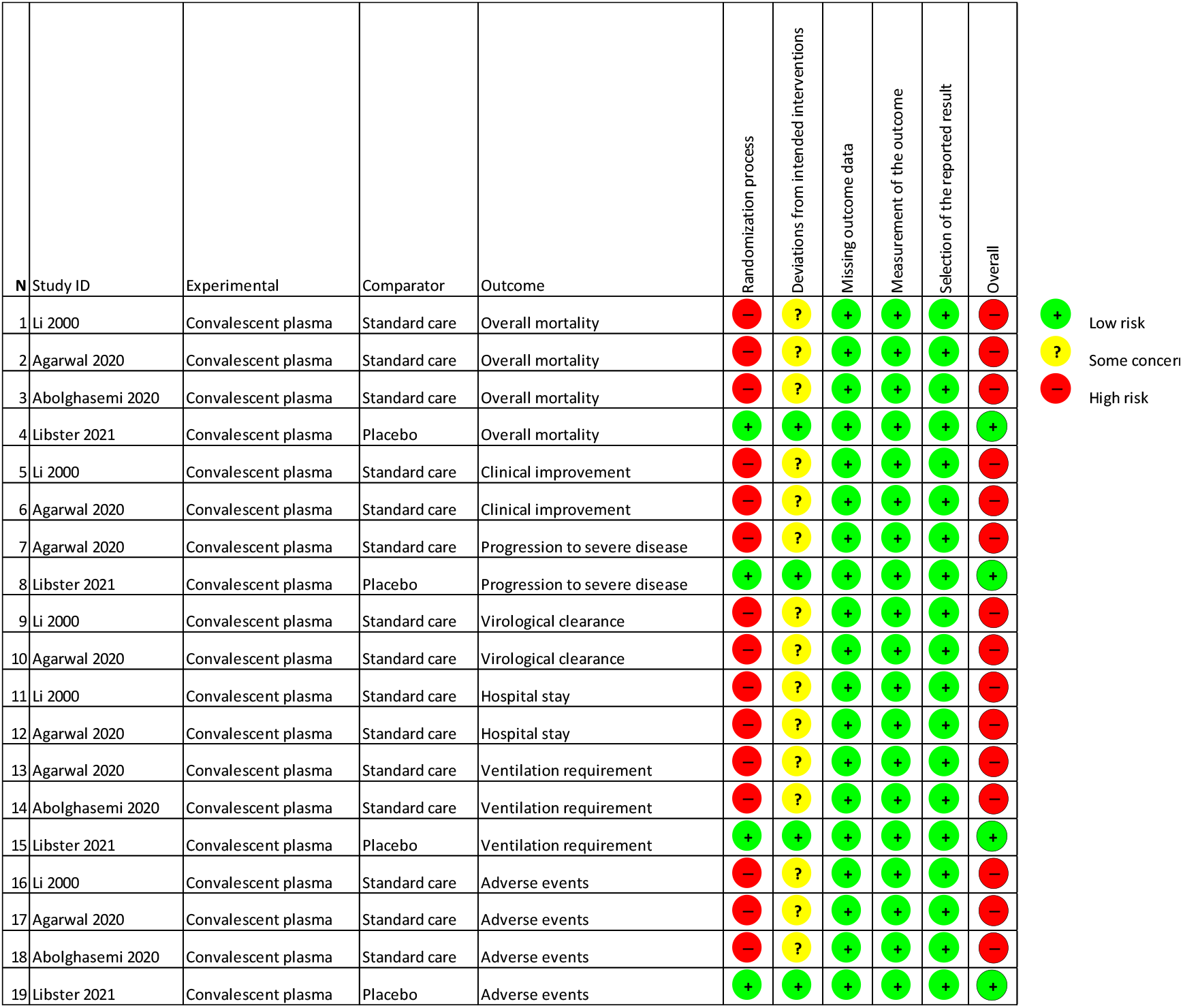

**Table 2.** Excluded studies with reasons

| Reference | Author | Reason for excluding |
| --- | --- | --- |
| (1) | Salazar | single arm |
| (2) | Wang | no control group |
| (3) | Skendros | in vitro study |
| (4) | Choudhuru | protocol |
| (5) | Olivares-Gazca | no control group |
| (6) | Teofili | protocol |
| (7) | Bradfute | single arm |
| (8) | Mayer | data science study |
| (9) | Mo | not trial |
| (10) | Rodríguez-Rubio | protocol |
| (11) | Ragnesola | single arm |
| (12) | Johansson | another drug |
| (13) | Li | duplicated |
| (14) | Agarwal | duplicated |
| (15) | Kumar | not trial |
| (16) | Omrani | no control group |
| (17) | Moore | case report |
| (18) | Khamis | case series |
| (19) | Meo | not trial |
| (20) | Snawerdt | not trial |
| (21) | Moniuszko-Malinowska | case report |
| (22) | Ibrahim | single arm |
| (23) | Liu | retrospective |
| (24) | González | Observational |
| (25) | Liu | case report |
| (13) | Li | duplicated |
| (26) | Saverino | not trial |
| (27) | Chen | not trial |
| (28) | Korper | protocol |
| (29) | Bobek | case report |
| (30) | Perotti | protocol |
| (31) | Shen | case report |
| (32) | Duan | case series |
| (14) | Agarwal | duplicated |
| (33) | Devos | protocol |
| (34) | Lou | another drug |
| (35) | Salazar | not trial |
| (36) | Su | not trial |
| (37) | Jorgensen | not trial |
| (38) | Blot | single arm |
| (39) | Dulipsingh | not trial |
| (40) | Dushiantan | another drug |
| (41) | Perreault | letter to editor |
| (42) | Wang | another drug |
| (43) | Lee | not trial |
| (44) | Erkurt | single arm |

